# Toscana Virus Infection Clinical Characterization

**DOI:** 10.1101/2024.07.31.24310805

**Authors:** Nazli Ayhan, Carole Eldin, Remi Charrel

## Abstract

**Background:** Toscana virus (TOSV) is a sandfly-borne phlebovirus causing central nervous system (CNS) infection in Mediterranean countries, during summer season. However, clinical aspects of the disease caused by this virus are poorly known by clinicians, so that its prevalence is probably underestimated due to a lack of diagnosis.

**Study design:** We gathered data from all available case series and retrospective studies identifying TOSV as the causative viral agent. The informations of age, sex, clinical characteristics, laboratory findings, imaging results and clinical outcomes of TOSV infection were recorded and analyzed.

**Results:** In our review a total of 95 articles including TOSV infections resulting in a total of 1,381 cases were analyzed. Our findings indicate, TOSV affects individuals across various age groups, with a median age of 44.45 years. A notable disparity in infection rates between genders, with men being significantly more likely to present symptoms due to TOSV than women, with a sex ratio of 2.0 (p<0.001). The clinical presentation of TOSV infection encompasses a range of symptoms, including fever, headache, retro-orbital pain, neurological and muscular manifestations with less common reports of cutaneous and gastrointestinal symptoms. To date, six fatalities have been attributed to TOSV infections, with a median age of 76 years.

Diagnostic evaluation of TOSV infections often involves the analysis of cerebrospinal fluid, where findings may include an elevated white blood cell count.

**Conclusions:** These findings underscore the diverse clinical manifestations of TOSV infections and highlight the importance of considering this pathogen in the differential diagnosis of patients presenting with acute febrile illness, especially in endemic regions. TOSV represents an emerging infectious threat that warrants inclusion in the diagnostic protocols for patients presenting with CNS, particularly within the Mediterranean basin or for those with recent travel history to endemic regions during warmer months when sandflies are actively circulating.

## Introduction

Toscana virus (TOSV) is classified within the *Phlebovirus toscanaense* species of the *Phenuiviridae* family within the *Bunyavirales* order. TOSV is recognized as a significant human pathogen prevalent in the Mediterranean region (1). Recent studies, including Medlock et al. (2) have documented a concerning increase in the population density of blood-feeding insects, along with their dissemination into territories previously considered free of sand flies. This expansion of the sand fly habitats substantially increases the risk of TOSV exposure among human population. Cases of TOSV infection have been reported in various Mediterranean countries such as; Italy, Spain, Portugal, France, Türkiye, Croatia, Greece, Algeria, Tunisia (3). Seroprevalence studies in both human and non-human vertebrates have revealed significant infection rates, with high prevalence reported in Italy (19.8%) (4), Türkiye (17.8%) (5), Greece (21%) (6), North Africa (22%-41%) (7); and the Balkans (37.5%) (8) indicating the widespread presence of TOSV across the Mediterranean basin. A recent retrospective study in Germany by Dersch et al. (2021) identified cases of TOSV-neuroinvasive disease in patients with meningoencephalitis with no recent travel history to endemic areas, suggesting a broader geographical spread than previously thought (9). TOSV is listed in the first three viral agents causing neurological infection at least in Italy, Spain and France together with enteroviruses and herpesviruses during warm season (10). However, the rare inclusion of TOSV in the diagnostic algorithm of nervous system infections (CNS) infections results in an important underestimation of the incidence (3).

The objective of our study was to review the available data about clinical characteristics of TOSV infections to raise the awareness of physicians about this emerging pathogen.

## Material and Method

### Study design

Relevant entries in global Web-based resources that comprise Scopus (http://www.scopus.com/), Web of Science (https://isiknowledge.com), and PubMed (https://pubmed.ncbi.nlm.nih.gov), Google scholar (https://scholar.google.com.tr/) were searched. Database investigations were performed using the keywords “bunyavirus”, “phlebovirus,”, “Toscana virus”, “TOSV”, “Toscana virus case report”, ”nervous system infection”. Reports unrelated to TOSV infection were omitted, as well as conference reports with recurring data in publications. The references cited in each report were examined for further publications, which were included in the analysis (Figure 1).

**Figure 1.**
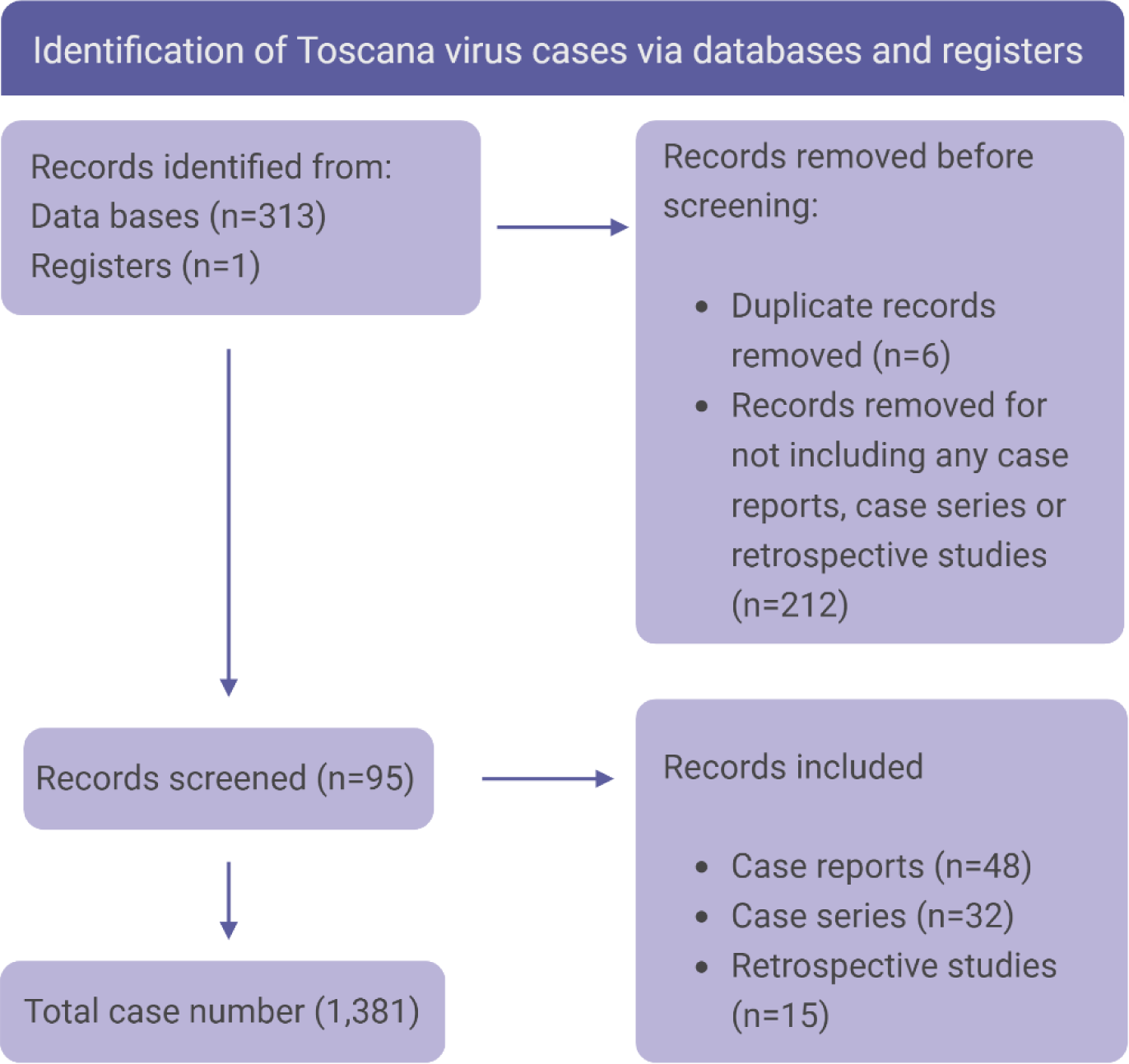
Toscana virus local and imported case distribution map.

### Statistical Analysis

All variables (described in Table 1) were screened using a logistic regression univariate analysis to check for statistically significant associations of demographic characters with TOSV in SPSS version 24.

**Table 1.**
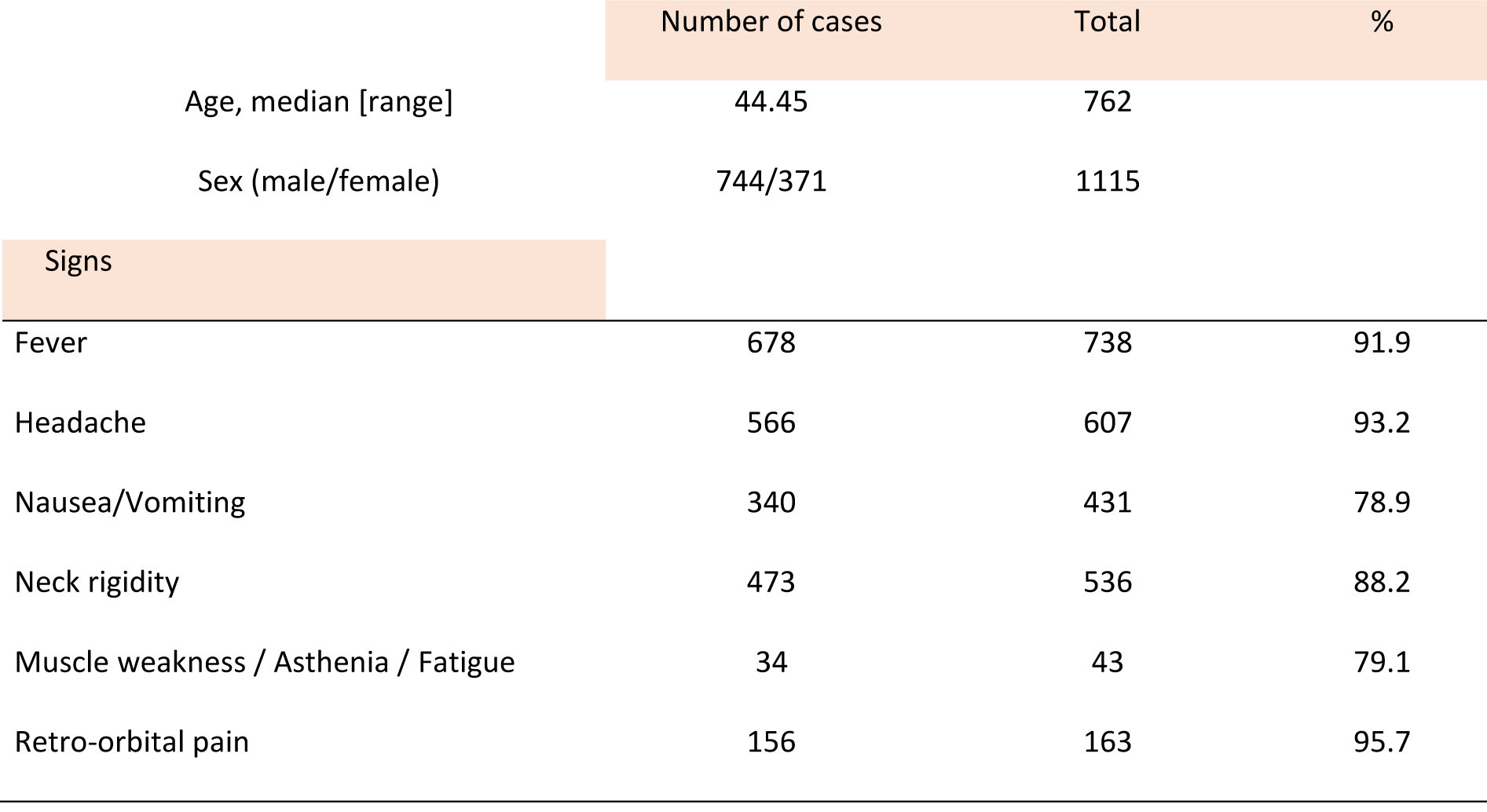
Demographic characteristics and non-specific clinical signs.

## Results

### Demographic and geographic characteristics

We conducted an inventory of 95 articles including 48 case reports, 32 case series and 15 retrospective studies, documenting a total of 1,381 cases of TOSV infection cases between 1985 and 2023. Age and sex demographics were obtained for 762 and 1,115 patients respectively (Table 1). Significantly more male patients were reported with TOSV infection compared to female patients (744 males to 371 females; p<0.001).

TOSV infections were recorded in 12 countries: with the majority of cases originating from Italy (n=1064), followed by Spain (n=76), Greece (n=45), Türkiye and Tunisia each with (n=31), France (n=27), Algeria (n=23), and lesser numbers from Croatia (n=12), Portugal (n=12), Romania (n=8), Bosnia & Herzegovina (n=7), Germany (n=4) and Israel (n=1). A total of 40 travelers returning from Mediterranean countries were reported, of which 30 had visited Italy, including both mainland and islands (Figure 2).

**Figure 2.**
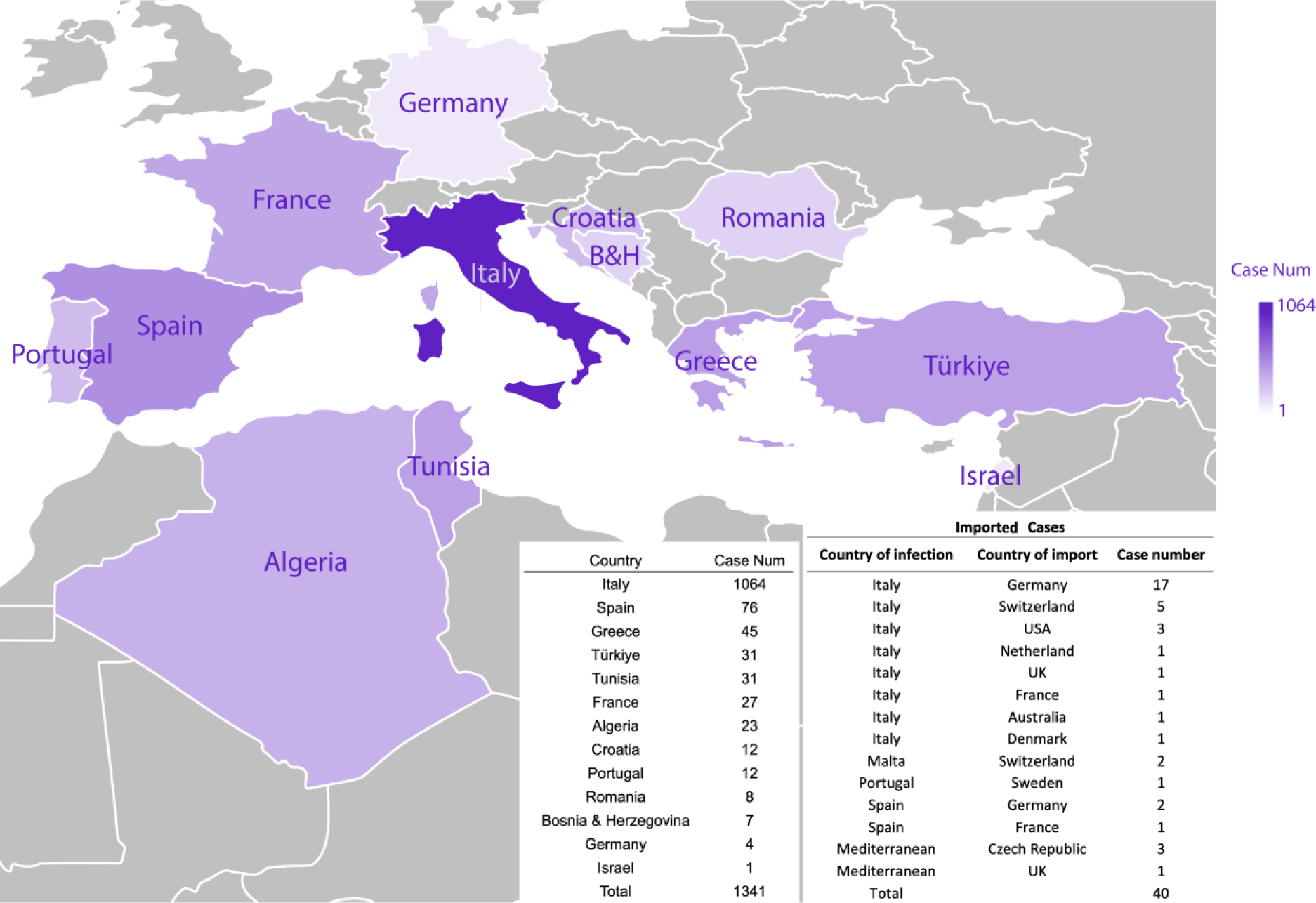
Toscana virus local and imported case distribution map.

TOSV infections were reported across a wide range of age groups, with the youngest patient being between 0-5 year old (11) and the oldest between 90-95 year old (12); the median age of patients was 44.4 years. The disease manifested in 82 pediatric patients under 15 years of age, as documented in nine studies (11–19). Notably, the prevalence of TOSV infection in children was significantly lower, at 5.9%.

### Clinical characteristics

#### Typical manifestations

Retro-orbital pain was the most common symptom (156/163, 95.7%). Then headache was observed in 93.2% of cases (566/607) sometimes reported as the “worst headache of their life”. Fever was described in 91.9% (678/738) of cases with an abrupt onset and temperature ranging from 38°C to 39.5°C. Neck rigidity was reported in 88.2% (473/536). Other non-specific signs such as nausea/vomiting, muscle weakness, asthenia and fatigue were described in almost 80% of cases (Table 1).

#### Neurological manifestations

Of the 644 documented cases, 519 patients (representing 80.6%) presented with meningitis or aseptic meningitis, while 111 patients presented with either pure encephalitis or meningo-encephalitis (Table 2); myelitis was very rarely reported. Until recently, a total of 49 cases were recorded as meningitis/meningo-encephalitis. More recently, Mellace et al. (2022) identified 331 cases featuring neurological symptoms; however, the distinction between meningitis and encephalitis/meningoencephalitis was not clearly defined.. As a consequence, although central manifestations are not unusual, pure meningitis is the most prominent neurological manifestation of TOSV infections.

**Table 2.**
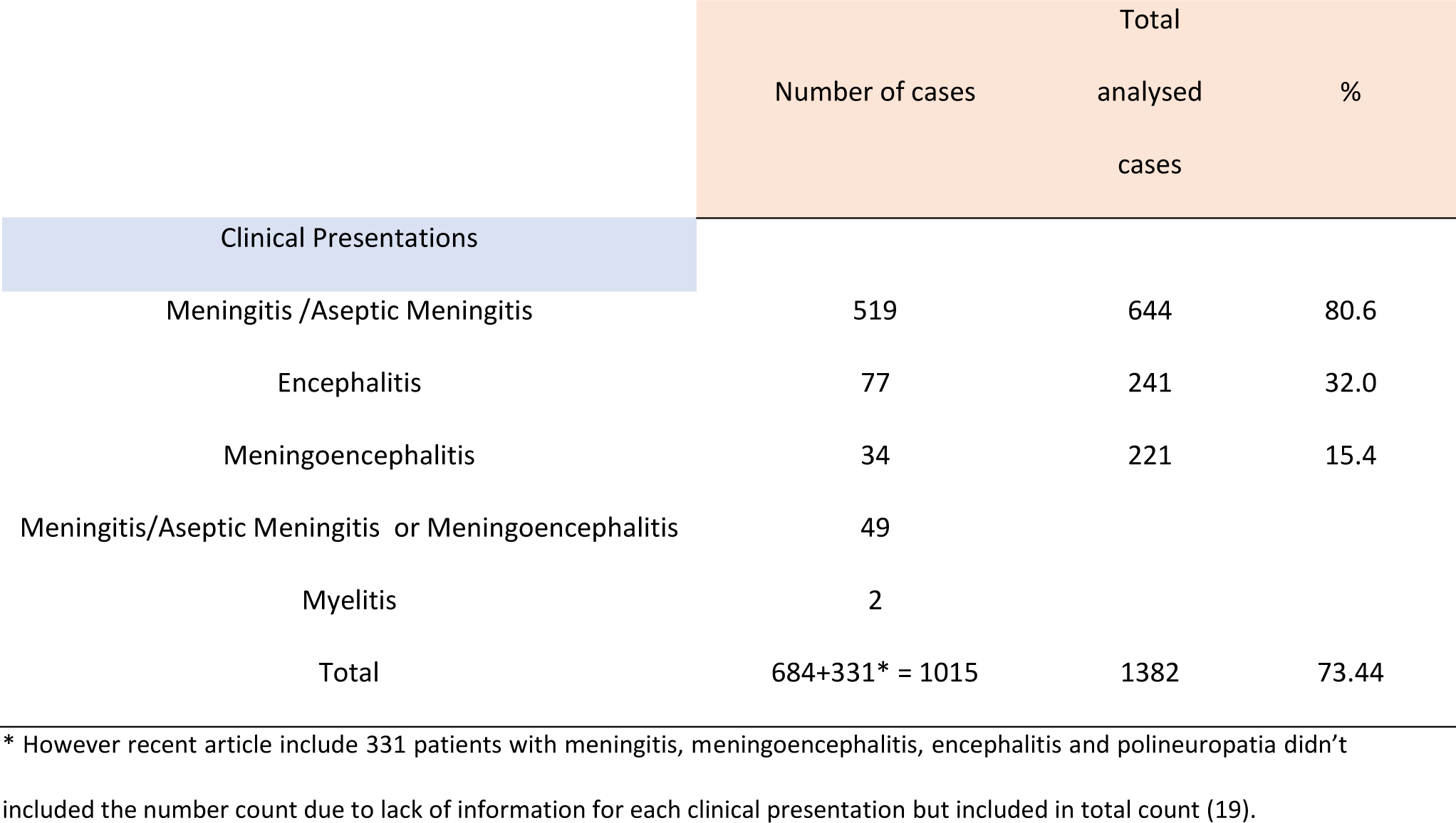
TOSV neurological manifestations.

Three cases of hydrocephalus were documented in young patients (range between 15-25 year-old); in all cases, hydrocephalus was observed as a complication following viral meningoencephalitis (20,21).

Specific neurological manifestations were categorized based on their involvement of either the central or peripheral nervous system, as detailed in Table 3.

**Table 3.**
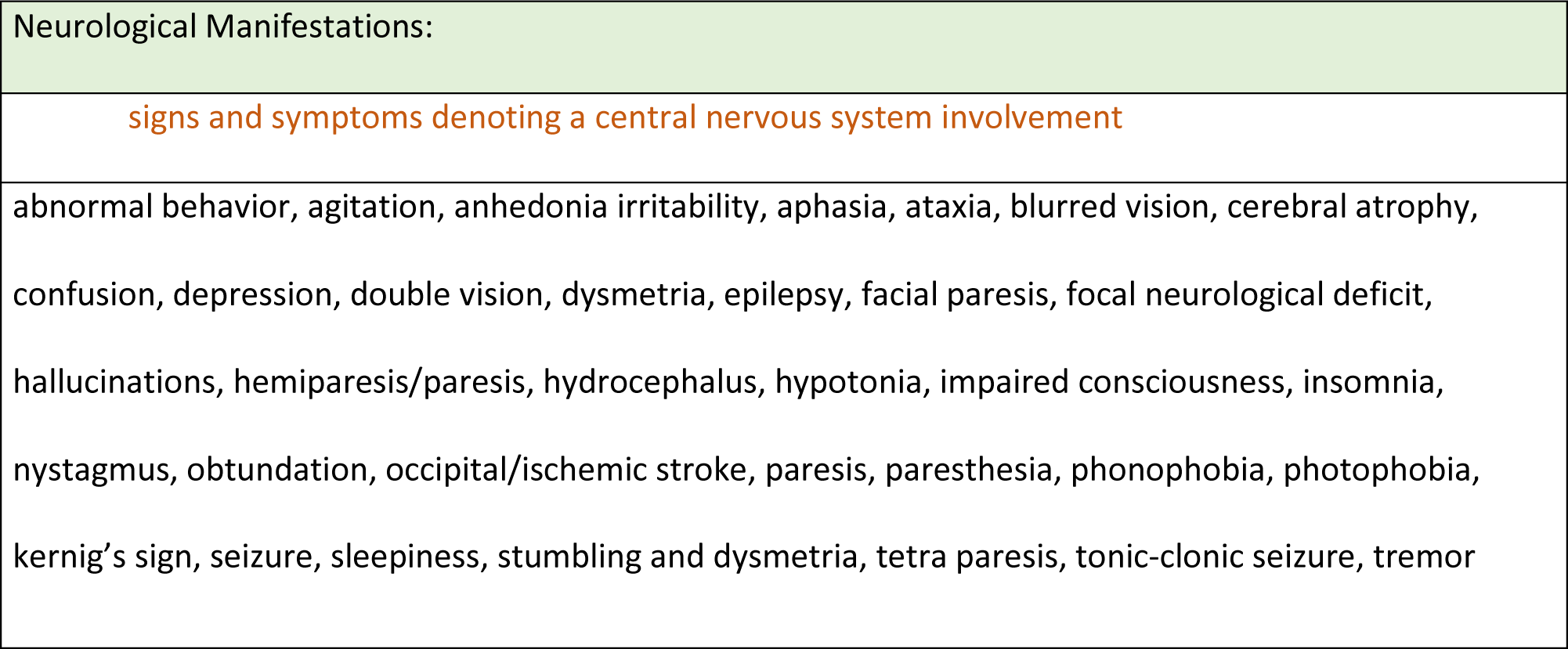
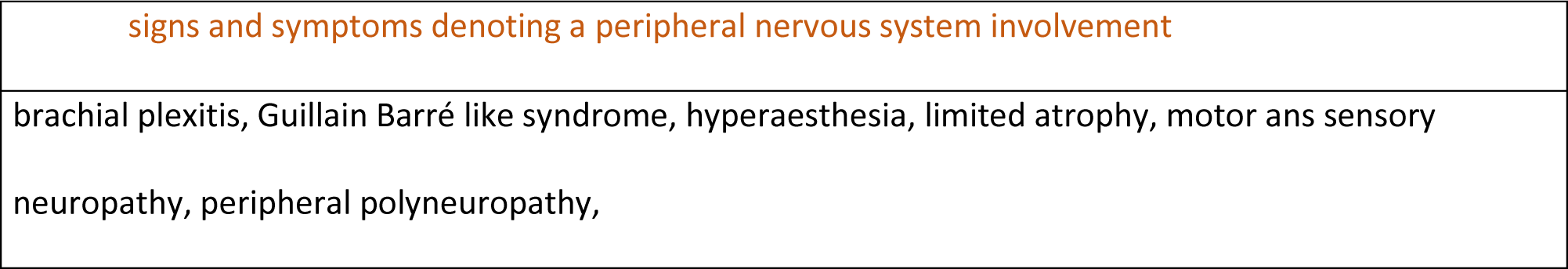
TOSV neurological signs and symptoms.

|  |
| --- |
| Neurological Manifestations: |
| signs and symptoms denoting a central nervous system involvement |
| abnormal behavior, agitation, anhedonia irritability, aphasia, ataxia, blurred vision, cerebral atrophy, confusion, depression, double vision, dysmetria, epilepsy, facial paresis, focal neurological deficit, hallucinations, hemiparesis/paresis, hydrocephalus, hypotonia, impaired consciousness, insomnia, nystagmus, obtundation, occipital/ischemic stroke, paresis, paresthesia, phonophobia, photophobia, kernig's sign, seizure, sleepiness, stumbling and dysmetria, tetra paresis, tonic-clonic seizure, tremor |
| signs and symptoms denoting a peripheral nervous system involvement |
| brachial plexitis, Guillain Barré like syndrome, hyperaesthesia, limited atrophy, motor and sensory neuropathy, peripheral polyneuropathy, |

**Table 4.** Number of TOSV cases with muscular, ocular, gastro-intestinal, cutaneous and neurological manifestations.

|  | Number of cases | Total | % |
| --- | --- | --- | --- |
| Muscular Manifestations | 51 | 211 | 24,2 |
| Myalgia / Arthralgia | 51 | 211 | 24,2 |
| Ocular Manifestations | 157 | 164 | 95,7 |
| Retroorbital pain or pressure | 156 | 163 | 95,7 |
| Gastro-intestinal Manifestations | 16 | 43 | 37,2 |
| Gastroenteritis | 7 | 16 | 43,8 |
| Cutaneous Manifestations | 15 | 38 | 39,5 |
| Rash | 11 | 32 | 34,4 |
| Neurological Manifestations | 569 | 712 | 79.9 |
| Central Nervous System Manifestations | 560 | 683 | 82 |
| Peripheral Nervous System Manifestations | 14 | 26 | 53,8 |

Out of 712 patient, 569 exhibited at least one neurological signs or symptoms listed in Table3. Among central neurological manifestations, Kernig’s sign was the most prevalent, observed in 135 cases, followed by decreased consciousness and photophobia (n=41), facial or leg hemiparesis /paresis (n=17) and confusion (n=16). Speech impairmentwas reported in eight patients(22–28) while six patients experienced hearing impairment, characterized by bilateral deafness that persisted beyond the acute phase and 6 patients reported hearing impairment (29–32). Additionally, changes in personality, sexual and social disinhibition, aggressiveness, and other abnormal behaviors were also documented (33,34) (Table2). The presence of symptoms signing a peripheral neuropathy was observed in 14 cases of which 10 presented with Guillain-Barre like syndrome (35–37).

#### Ocular manifestations

Retro-orbital pain or pressure were very frequently observed, documented in 156 out of 163 cases. In contrast, conjunctivitis was reported only in a single case (38) (Table4).

#### Gastro-intestinal manifestations

Gastro-intestinal manifestations consisting of gastroenteritis, abdominal pain, dysphagia or diarrhea were reported rarely (16/43). (20,37,39–41) (Table4).

#### Cutaneous manifestations

Dermatological symptoms including petechiae, rash, exanthema, and febrile erythema were reported in 15 out of 38 cases. Rash was the most frequently observed symptom, noted in 11 cases (20,29,39,41–45) (46) (Table4).

#### Muscular manifestations

The musculoskeletal symptoms observed included cramps, myalgia/arthralgia, myositis/fasciitis, and muscle stiffness. Among the 51 patients who exhibited these symptoms, all presented with myalgia and/or arthralgia (12,18,22,35,38,44,45,47–53) (Table4).

#### Testicular manifestations

Testicular manifestations, including epididymo-orchitis, testicular pain, and swelling, were reported in five patients (20,39,41,54,55) (Table 5). These symptoms were significantly more frequent (p-value <0.001) among younger patients, with a median age of 27 years (range 16-44) (Table 5). Recently, TOSV RNA was detected in the seminal fluid of a patient in the age range of 20-25 year old until day 59 after infection without any testicular manifestations (28).

**Table 5.** TOSV infection cases with testicular manifestations.

| Characteristic / References | Age range / Age<br>median [range] (n) | Fever | Headache | Neck rigidity /<br>pain | Nausea /<br>vomiting |
| --- | --- | --- | --- | --- | --- |
| Echevarria et al. 2003 | 25-29 | NR* | NR* | NR* | NR* |
| Baldelli et al. 2004 | 15-19 | 1 | 1 | 1 | 1 |
| Zanelli et al. 2013 | 25-29 | 1 | 1 | - | - |
| Tschumi et al. 2019 | 20-24 | 1 | 1 | - | - |
| Mascitti et al. 2020 | 40-44 | 1 | 1 | - | - |
|  | 27 [16-44] (5) | 4 | 4 | 1 | 1 |

\*, NR, not reported

#### Severe cases and Mortality

Although TOSV infections are frequently associated with severe clinical manifestations, the vast majority of patients recover completely. Nonetheless, six fatal cases have been reported across two studies: the first fatality occurred in Italy (31); and five subsequent fatalities were reported in Romania (31,49). The overall the mortality rate is 0.43% with deceased patients having a median age of 76 years-old (range 70-91). Age was found to have a statistically significant impact on mortality (*p*-value <0.001). In addition, five out of six fatal cases had comorbidities such as hypertension and diabetes (Table6).

There were nine cases of severe TOSV infections leading to coma (20,31,49,56). Seven of these were elder patients with median age of 79, six of whom had comorbidities such as diabetes and hypertension and two patients were two siblings with the age range between 15 to 20, previously mentioned as having developed hydrocephalus following infection.

#### Laboratory characteristics

Cerebrospinal fluid (CSF) analysis showed elevated white blood cell (WBC) levels in 22 cases. Lymphocytic predominance ratio was high in 82% of the patients (47 out of 57) (>50%). CSF samples showed high protein levels in 102 patients. Mildly elevated glucose levels have been showed in 179 patients (Table 7).

**Table 6.** TOSV infection lethal cases.

| Reference | Nb of case | Sex | Age Range | Country | Comorbidities | Period | In-hospital length of stay (days) |
| --- | --- | --- | --- | --- | --- | --- | --- |
| Bartels et al. 2012 | 1 | male | 70-74 | Italy | - | - | 14 |
| Popescu et al. 2021 | 1 | male | 90-94 | Romania | Hypertension, congestive heart failure, ischemic heart disease, stroke sequelae | June | 10 |
|  | 1 | female | 65-69 | Romania | Hypertension, diabetes mellitus, obesity, NonHodgkin lymphoma | August | 4 |
|  | 1 | male | 75-79 | Romania | Diabetes mellitus type II, Ischemic heart disease | August | 19 |
|  | 1 | male | 70-74 | Romania | Diabetes mellitus type II, Ischemic heart disease, Atrial fibrillation, Congestive heart failure, Diabetic polyneuropathy and arteriopathy | August | 4 |
|  | 1 | female | 85-89 | Romania | Hypertension, Stroke sequelae, Chronic renal failure | August | 6 |

**Table 7.** TOSV laboratory characteristics.

| Laboratory Findings | Mean (n) [range] | Normal range/percentage |
| --- | --- | --- |
| CSF protein level (mg/dL) | 56.34 (77) [11-757] | 15-45 |
| CSF WBC (cells/mm3) | 432.50 (22) [6-3500] | 0-5 |
| CSF Lymphocytes ratio (%) | 73.40% (57) [18%-100%] | 50% |
| CSF Glucose (mg/dL) | 63.21 (60) [29-132] | >40 |

**Table 8.** TOSV electro-physiology analysis and radiology findings.

| Electro-physiology analysis and<br>radiology findings | Number of abnormal cases | Total |
| --- | --- | --- |
| EEG | 43 | 82 |
| MRI | 11 | 30 |
| CT | 3 | 30 |
| Chest X-ray | 1 | 8 |
| ECG | 0 | 3 |

#### Electro-physiology analysis

The electroencephalogram (EEG) showed abnormal results in 43 patients (11,23,31,56–62) with non-specific abnormalities, occasional spikes and slow waves (58).

#### Imaging and Radiological findings and electrocardiogram tests

When available, magnetic resonance imaging (MRI) results showed diffuse encephalopathy, diffuse atrophy abnormalities, diffuse bilateral asymmetric myositis or scattered punctuate T2-hyperintense non-enhancing white matter lesions in 11 patients (21,27,45,53,63). Cerebral atrophy and/or occipital stroke sequelae were detected in 2 patients with computerized tomography (CT) scan (49).

Few patients were examined by with electrocardiogram (ECG) and chest X-ray. Chest X-ray demonstrated an opacity in the right middle lobe of the lung of one patient (29). No abnormalities were recorded with ECG after TOSV infection (Table8).

## Discussion

Infections caused by TOSV exhibit a high diversity of clinical manifestations, none of them being neither pathognomonic nor highly specific. It is crucial to provide a comprehensive analysis of the observed signs in order to improve the clinical orientation for physicians seeing both residents and visitors in the Mediterranean regions during the warm season when TOSV can be transmitted by sand flies (Figure1). Unlike other neuroinvasive viruses such as herpes viruses and enteroviruses, TOSV infections occur exclusively during the period of activity of phlebotomine sand flies, typically from March to November in the Mediterranean, with slight variations depending on local climate conditions (64).

From 1985 to2023, a total of 1,381 cases were reported from 12 different countries of which the majority (1,064 cases, 77%) occurred in Italy. The annual incidence rate of cases fluctuates considerably in relation with complex biology of vectors, hosts and environmental factors. TOSV infections have been documented across a broad age spectrum (median age 44.45 years, ranging from 0 to 95 years old); however, age itself is not indicative of etiology, though most cases occur between 20 and 60 years old. Using data collected in travelers returning to their homeland, the median incubation period was calculated at 12 days (10-14 days) for the neuroinvasive forms (65). Whether the same incubation period applies for milder cases remains unexplored.

TOSV symptomatic infections are observed twice as frequently in males as in females. The reasons for this gender disparity in symptomatology could be related to differences in immune response, viral susceptibility, or possibly due to variations in exposure factors and the prevalence of high-risk behaviors. However, similar seroprevalence rates between males and females suggest that behavioral factors alone may not explain this difference (66). Interestingly, the same tendency is also observed for WNV infections (67).

Both length of hospitalization and symptoms duration vary significantly based on the severity of the case. The longest hospitalization was 60 days (68) and the longest symptoms duration was 28 days (52,69). The letality rate of TOSV infections is 0.43% which is lower than other neuroinvasive arboviruses such as WNV, Japanese encephalitis virus (JEV) or tick-borne encephalitis virus (TBEV). All the fatal cases were older than 60 year-old; five of the six fatal cases had comorbidities such as hypertension and/or diabetes mellitus.

Severe neuroinvasive forms, characterized by encephalitis or meningo-encephalitis, are reported in 15.8 to 32.0% of neurological cases, the latter representing almost 80% of 712 studied cases. Since TOSV cases reported in the literature are the most severe ones, neuroinvasive forms are certainly overrepresented. Common forms are frequent although they are likely to remain either undetected or unpublished (70); those forms are associated with unspecific clinical manifestations such as fever, headache, nausea, vomiting, retro-orbital pain and muscle weakness.

Severe long-lasting or permanent neurological sequelae such as consciousness impairment, hydrocephalus, ischemic stroke and aphasia (20,49,68,71,72) were described. In contrast with other arboviruses, severe neurological forms are not more frequent in the elderly (19). However, the lower incidence rate of TOSV infections in the pediatric group (5.9%) can either be explained by a lower exposure and/or by a reporting bias, or by biological factors such as immune response and/or viral susceptibility.

Among system specific manifestations, CNS signs are the most frequently observed with 79.9% of cases followed by ocular and muscular manifestations. Peripheral neurological manifestations, gastro- intestinal and cutaneous manifestations were reported rarely. Additionally, a unique case involved a patient with gastrointestinal manifestations who was also co-infected with West Nile virus (37). Although testicular manifestations are noteworthy, they have been documented in only five patients. To date, aside from the Zika virus (ZIKV), no other arbovirus has been identified as a causative agent for testicular manifestations. Recently, the presence and persistence of TOSV RNA in seminal fluid has been demonstrated without evidence for sexual transmission (28).

In most of cases presenting with clinical picture justifying CSF collection, CSF was clear and colorless. The CSF formula showed elevated WBC levels in 22 patients and lymphocytic meningitis in 58 patients. Mildly elevated protein and glucose levels were described in 102 patients and 179 patients respectively. Hypoglycorachia was never reported. CSF parameters were available only for a subset of the reviewed cases.

Electro-physiology analysis and radiology findings showed abnormalities in EEG or in MRI for 56 cases (4%). CT scan and chest X-ray were performed occasionally and abnormalities were rarely recorded. No abnormalities were recorded for ECG which was performed only for three patients (58,73,74) In almost all cases, the recovery is complete without persisting functional sequelae. However, there are a few documented cases where individuals have experienced complicated forms of meningitis and/or encephalitis with lingering effects attributed to TOSV. These effects include impaired cognitive functions and altered social and sexual behaviors (60,68) Other neuroinvasive arboviruses, such as WNV, JEV or TBEV, cause long-lasting or permanent sequelae (75).

Finally, most TOSV infections remain undiagnosed, as TOSV is not included in the list of pathogens to be screened in patients presenting with febrile illness and/or neurological manifestations in areas where the virus is endemic. The only exception is Italy where it is recommended to include TOSV in the panel of viruses to be tested during summertime for suspect cases. This is likely why most cases are reported in Italy besides the fact that TOSV has been discovered in Italy and that physicians are much more aware of its existence than in other at risk countries. This oversight is highlighted by numerous retrospective studies, which suggest that many cases of TOSV infection are classified as “infections due to unknown pathogenic agents“ due to the absence of specific laboratory screening for TOSV (35,51,76,77). Nevertheless, most TOSV cases exhibit nonspecific signs or symptoms with short duration in non-severe cases.

Together, the combination of lack of awareness of physician with absence of recommendations to screen suspect cases using specific laboratory tests has and continue to greatly contribute to the underestimation of TOSV cases despite its public health importance.

## Data Availability

All data produced in the present study are available upon reasonable request to the authors.

## Acknowledgement

This study was funded by European Commission grant 101057690 and UKRI grants 10038150 and 10039289, and is catalogued by the CLIMOS Scientific Committee as CLIMOS number XXXX* (http://www.climos-project.eu). The contents of this publication are the sole responsibility of the authors and do not necessarily reflect the views of the European Commission, the Health, and Digital Executive Agency, or UKRI. Neither the European Union nor granting authority nor UKRI can be held responsible. The funders had no role in study design, data collection and analysis, decision to publish, or preparation of the manuscript. For the purposes of Open Access, the authors have applied a CC BY [option: CC BY-ND] public copyright license to any Author Accepted Manuscript version arising from this submission. CLIMOS forms part of the Climate Change and Health Cluster comprising six Horizon Europe projects: BlueAdapt, CATALYSE, CLIMOS, HIGH Horizons, IDAlert, and TRIGGER.

*The number will be added after publication.

